# SARS-CoV-2 Seroepidemiology in Children and Adolescents

**DOI:** 10.1101/2021.01.28.21250466

**Authors:** Rebecca E Levorson, Erica Christian, Brett Hunter, Jasdeep Sayal, Jiayang Sun, Scott A Bruce, Stephanie Garofalo, Matthew Southerland, Svetlana Ho, Shira Levy, Christopher Defillipi, Lilian Peake, Frederick C Place, Suchitra K Hourigan

## Abstract

**Objectives:** Pediatric SARS-CoV-2 data remain limited and seropositivity rates in children were reported as <1% early in the pandemic. Seroepidemiologic evaluation of SARS-CoV-2 in children in a major metropolitan region of the United States was performed.

**Methods:** Children and adolescents ≤19 years were enrolled in a cross-sectional, observational study of SARS-CoV-2 seroprevalence from July-October 2020 in Northern Virginia, United States. Demographic, health, and COVID-19 exposure information was collected, and blood was analyzed for SARS-CoV-2 spike protein total antibody. Risk factors associated with SARS-CoV-2 seropositivity were analyzed. Orthogonal antibody testing was performed, and samples were evaluated for responses to different antigens.

**Results:** In 1038 children, the anti-SARS-CoV-2 total antibody positivity rate was 8.5%. After multivariate logistic regression, significant risk factors included Hispanic ethnicity, public or absent insurance, a history of COVID-19 symptoms, exposure to person with COVID-19, a household member positive for SARS-CoV-2 and multi-family or apartment dwelling without a private entrance. 66% of seropositive children had no symptoms of COVID-19. Orthogonal antibody testing with a receptor binding domain specific antigen revealed a high concordance of 80.5%. Children also demonstrated a robust immune response to the nucleocapsid antigen.

**Conclusions:** A much higher burden of SARS-CoV-2 infection, as determined by seropositivity, was found in children than previously reported; this was also higher compared to adults in the same region at a similar time. Contrary to prior reports, we determined children shoulder a significant burden of COVID-19 infection. The role of children’s disease transmission must be considered in COVID-19 mitigation strategies including vaccination.

**Article Summary:** 8.5% of children had SARS-CoV-2 antibodies in Fall 2020, double the adult rate. The role of pediatric infection is important to consider in mitigation strategies.

**What’s Known on This Subject:** SARS-CoV-2 pediatric seroepidemiologic data is limited. Reported viral rates underestimate the burden of infection in children due to mild or asymptomatic disease. Limited cohorts of children suggest low seropositivity rates compared to adults.

**What This Study Adds:** US children in the largest SARS-CoV-2 seroepidemiology study to date had double the rate of antibodies compared to adults. Most children were asymptomatic. Risk factors include age, ethnicity and living conditions. Most children made antibodies to different antigens of SARS-CoV-2.

## Introduction

Current epidemiologic data in the United States (US) and globally indicating sustained community transmission of SARS-CoV-2 is indisputable. The extent of SARS-CoV-2 community transmission remains incompletely understood, particularly regarding children. Initially, our understanding was dependent upon aggregate molecular testing for acute infection and largely missed mild and asymptomatic infections. Based on this data, early reports misleadingly suggested that children were largely spared infection from SARS-CoV-2 [1,2].

Seroprevalence estimates can be useful to understand the true cumulative incidence in a given population. Antibody seropositivity provides data about asymptomatic or subclinical infections that would not be detected by surveillance relying on interactions with the health care system. Limited pediatric seroprevalence data exist for SARS-CoV-2. Following the first pandemic wave in Spain, up to a 6.2% seropositivity was reported late April/early May 2020, with only 3.8% of children aged 0-19 years being seropositive [3]. Antibody positivity in a smaller population-based study in Switzerland in mid-May 2020 was lowest in children 0-9 years at 0.8%, compared with adults aged 20-49, at 9.9% [4]. Similarly, in Germany a low seropositivity rate of 0.6% was reported in children aged 1-10 years [5],

An investigation in spring 2020 evaluated children for SARS-CoV-2 antibodies 8-10 weeks after a large school outbreak in Santiago, Chile. Anti-SARS-CoV-2 antibodies were present in 10% of children, whereas 17% of adult staff were seropositive. Forty percent of seropositive children were categorized as asymptomatic [6]. Outside of this outbreak associated investigation, there have been no large pediatric focused seroprevalence studies in the Americas.

Compared to adults, preliminary evidence in children suggests a distinct antibody response to SARS-CoV-2 infection, with a reduced breadth of anti-SARS-CoV-2 antibodies, specifically a reduction in anti-nucleocapsid antibody production [7]. Furthermore, the possibility has been raised that children may have a higher prevalence of antibodies against the S2 unit of the SARS-CoV-2 spike protein, which may be partially due to cross-reactivity to conserved epitopes in benign seasonal human coronaviruses [8].

As there is extremely limited data regarding the burden of SARS-CoV-2 infection in children, especially in the US, the primary objective of this study was to determine SARS-CoV-2 seroprevalence in children residing in Northern Virginia in the US. Northern Virginia is the most populous and urbanized region of Virginia and is geographically linked to the Washington, D.C. metropolitan area. Secondary objectives included identification of risk factors associated with SARS-CoV-2 seropositivity in children, orthogonal antibody testing to assess the accuracy of antibody testing in children and evaluation of immunologic responses to different antigens of SARS-CoV-2 in children.

## Methods

Children and adolescents were enrolled in this cross-sectional, observational study over a 10-week period, from July 31 to October 13, 2020. Verbal informed consent was obtained from parents and adolescents ≥18 years. The Inova Health Systems Human Research Protection Office waived ethics approval as per CFR 45 46.102, as this project was a public health surveillance activity, funded by the Virginia Department of Health (VDH). During the study period, public schools in Northern Virginia were closed to in-person learning although, some childcare facilities remained open. An indoor mask mandate outside the home was in effect; however, this was only for children ≥10 years and adults.

Subjects were recruited from three settings in Northern Virginia: 1) non-emergency health care settings (including pediatric primary care offices, pediatric specialty clinics and pre-surgical areas for elective procedures); 2) self-referral in response to advertisement of the study, and 3) the Inova Children’s Hospital Pediatric Emergency Department and limited enrollment from inpatient units. Inclusion criteria were ≤19 years (the age range defined as a child for COVID-19 reporting by the state of Virginia) and residence in Virginia. Only one individual per household was permitted to enroll in the study. Exclusion criteria included receipt of immunoglobulin therapy within the past 11 months, including intravenous immunoglobulin, whole blood, fresh frozen plasma, experimental therapy with convalescent plasma, or other blood product transfusions, except for packed red blood cells and platelets.

Enrolled participants completed a questionnaire collecting demographic, health and potential COVID-19 exposure information. Blood was collected by venipuncture, either for the sole purpose of the study or in conjunction with another clinical blood draw. Blood was tested using the US Food and Drug Administration emergency use authorized Ortho Clinical Diagnostics VITROS Immunodiagnostic Products Anti-SARS-CoV-2 Total test (Ortho Clinical) performed on the Vitros 3600 system (Ortho Clinical Diagnostics, Raritan, NJ) to detect total antibody (IgG, IgA and IgM) responses against the SARS-CoV-2 spike protein. This assay has a reported clinical specificity of 100% (95% CI: 99.1-100%) [9]. Samples were documented as reactive (≥1.00 S/Co) or non-reactive (<1.00 S/Co) for anti-SARS-CoV-2.

Orthogonal antibody testing was performed on reactive samples as the estimated prevalence prior to the study was 1%, based on very limited data at the time.^4^ Orthogonal serologic testing was recommended by the CDC for populations with a low pre-test probability, including low or unknown prevalence of disease [10]. In such conditions, an initial antibody assay is performed, and then on all positive tests, a secondary assay with a different or more focused target is performed. In this study, the Ortho Clinical assay (targeting the total protein - S1 and S2 subunits) was used as the primary assay, followed by the Siemens SARS-CoV-2 IgG assay solely targeting the S1 unit receptor binding domain (RBD) (Siemens Healthineers, Erlangen, Germany). Additionally, to assess the antibody response in children to different SARS-CoV-2 antigens, all initially reactive samples were tested using the Abbot nucleocapsid IgG antibody assay (Abbott Laboratories, Chicago, IL). Furthermore, a random 10% sample of the initial non-reactive samples on the Ortho Clinical assay were analyzed on the Siemens (RBD) and Abbot (nucleocapsid) IgG assays.

Sample size justification was based on early estimates suggesting a SARS-CoV-2 seroprevalence in children of approximately 1% [4]. Thus, in order to estimate the seropositivity in demographic subgroups while considering the clustering effect due to testing at different enrollment sites, 80% Wilson score intervals were used to determine a total sample size of at least 1,000 children.

Statistical analysis methods included both comprehensive descriptive statistics and statistical modeling. Positive frequency counts were used to estimate both the overall and subgroup prevalence rates to delineate the effects of different factors. The corresponding Wilson score intervals provided the uncertainty quantification of the prevalence estimates. Graphical diagnostics and chi-squared tests of independence were used to determine whether selected covariates were correlated. Univariate and multivariate logistic regression analyses of antibody presence were used to explore potential effects from covariates. A final logistic regression model was chosen using a stepwise model selection. Odds ratios with corresponding confidence intervals were calculated for the coefficients in the model. The distribution of antibody titer levels across different factor levels was examined. Kruskal-Wallis tests were used to determine if these distributions differed across potential risk factors. Simple inter-rater reliability rates were calculated to assess orthogonal testing results. R version 4.0.3 was used to perform statistical analyses.

## Results

This study included 1038 children. Demographic and clinical data are shown in Table 1. All age groups between 0–19 years were well represented, and racial and ethnic diversity was reflective of the Northern Virginia populace.

**Table 1:** Demographic Information, Prevalence Rates, and 95% Wilson Score Intervals ^*^Indicates an unreliable estimate due to a small count.

|  |  |  |  |  |  |
| --- | --- | --- | --- | --- | --- |
| Variable | Category | Count | Estimated Prevalence Rate | 95% Wilson Score Intervals |  |
|  |  |  |  | Lower Bound | Upper Bound |
| Overall |  | 1038 | 8.5% | 6.9% | 10.3% |
| Gender | Male | 499 | 8.4% | 6.3% | 11.2% |
|  | Female | 536 | 8.6% | 6.5% | 11.3% |
|  | Not Disclosed | 3 | 0% * | 0% | 56.1% |
| Age Group | 0 – 5 years | 182 | 13.7% | 9.5% | 19.5% |
|  | 6 – 10 years | 241 | 7.5% | 4.8% | 11.5% |
|  | 11 – 15 years | 374 | 5.1% | 3.3% | 7.8% |
|  | 16 – 19 years | 241 | 10.8% | 7.5% | 15.3% |
| Race | White | 717 | 8.2% | 6.4% | 10.5% |
|  | Black or African American | 97 | 5.2% | 2.2% | 11.5% |
|  | Asian | 106 | 5.7% | 2.6% | 11.8% |
|  | American Indian or Alaska Native | 0 | --- | --- | --- |
|  | Native Hawaiian or Other Pacific Islander | 2 | 0% * | 0% | 65.8% |
|  | Other | 111 | 16.2% | 10.5% | 24.2% |
|  | Unknown/Declined | 5 | 0% * | 0% | 43.4% |
| Ethnicity | Not Hispanic or Latino | 829 | 4.0% | 2.8% | 5.5% |
|  | Hispanic or Latino | 207 | 26.6% | 21.0% | 33.0% |
|  | Unknown | 2 | 0% * | 0% | 65.8% |
| Insurance | Private | 834 | 5.3% | 4.0% | 7.0% |
|  | Medicaid | 125 | 21.6% | 15.3% | 29.6% |
|  | Medicare | 8 | 12.5% * | 2.2% | 47.1% |
|  | None or uninsured | 20 | 55.0% | 34.2% | 74.2% |
|  | Other | 3 | 33.3% * | 6.1% | 79.2% |
|  | Don't know | 4 | 25.0% * | 4.6% | 69.9% |
|  | Military | 44 | 6.8% | 2.3% | 18.2% |
| Existing Medical Comorbidity | No | 807 | 8.3% | 6.6% | 10.4% |
|  | Yes | 225 | 8.9% | 5.8% | 13.3% |
|  | Don't Know | 3 | 0% * | 0% | 56.1% |
|  | Missing | 3 | 33.3% | --- | --- |
| Symptoms | No | 774 | 7.2% | 5.6% | 9.3% |
|  | Yes | 255 | 11.8% | 0% | 65.8% |
|  | Unknown | 6 | 33.3% * | 9.7% | 70.0% |
|  | Missing | 3 | 0% | --- | --- |
| Dwelling | Single-family | 901 | 5.9% | 4.5% | 7.6% |
|  | Multi-family or apartment – No Private Entrance | 115 | 28.7% | 21.2% | 37.5% |
|  | Multi-family or apartment – Private Entrance | 16 | 6.3% | 1.1% | 28.3% |
|  | Other | 6 | 16.7% * | 3.0% | 56.4% |
| Household Member Exposure | Not Tested | 543 | 6.3% | 4.5% | 8.6% |
|  | Tested – Positive | 59 | 52.5% | 40.0% | 64.7% |
|  | Tested – Negative | 415 | 5.3% | 3.5% | 7.9% |
|  | Tested – No Results | 15 | 6.67% | 1.19% | 29.8% |
|  | Unknown if Tested | 4 | 0% * | 0% | 49.0% |
|  | Missing | 2 | 0% | --- | --- |
| COVID-19 Exposure | No | 911 | 5.6% | 4.3% | 7.3% |
|  | Yes | 106 | 33.0% | 24.8% | 42.4% |
|  | Unknown or Missing | 19 | 10.5% | 2.9% | 31.4% |
|  | Missing | 2 | 0% | --- | --- |
| Reason for Visit | Specifically for COVID-19 antibody testing | 504 | 4.6% | 3.1% | 6.8% |
|  | New or acute illness or injury | 193 | 14.0% | 9.8% | 19.6% |
|  | Prevention or well visit | 59 | 20.3% | 12.0% | 32.3% |
|  | Routine visit for a prior condition | 193 | 8.8% | 5.6% | 13.7% |
|  | Other | 84 | 10.1% | 5.4% | 18.1% |
| Traveled Out of the State | No | 395 | 13.4% | 10.4% | 17.1% |
|  | Yes | 642 | 5.5% | 3.9% | 7.5% |
|  | Don't Know | 1 | 0% * | 0% | 79.3% |
| Traveled Out of the Country | No | 1002 | 8.4% | 6.8% | 10.3% |
|  | Yes | 36 | 11.1% | 4.4% | 25.3% |
| Child Cared for outside of Home | No | 729 | 8.2% | 6.4% | 10.5% |
|  | Yes | 299 | 8.7% | 6.0% | 12.4% |
|  | Missing | 10 | 20.0% | --- | --- |
| Household Member Work Outside the Home | No work outside home | 383 | 6.3% | 4.2% | 9.2% |
|  | Essential | 530 | 8.7% | 6.6% | 11.4% |
|  | Unessential | 121 | 14.0% | 9.0% | 21.4% |
|  | Unknown if essential | 3 | 33.3% * | 6.1% | 79.2% |
|  | Missing | 1 | 0% | --- | --- |
| Enrollment Site | Pediatric Specialists Clinics | 444 | 4.7% | 3.1% | 7.1% |
|  | Pediatric Emergency Department | 175 | 14.3% | 9.9% | 20.2% |
|  | Pre surgical areas | 89 | 6.7% | 3.1% | 13.9% |
|  | Primary Care location 1 | 128 | 5.5% | 2.7% | 10.9% |
|  | Hematology/Oncology clinic | 57 | 17.5% | 9.8% | 29.4% |
|  | Primary Care location 2 | 64 | 25.0% | 16.0% | 36.8% |
|  | Pediatric Intensive Care Unit | 7 | 14.3% * | 2.6% | 51.3% |
|  | Pediatric inpatient units | 6 | 0% * | 0% | 39.0% |
|  | Endoscopy suite | 10 | 0% * | 0% | 27.8% |
|  | Primary Care location 3 | 49 | 4.1% | 1.1% | 13.7% |
|  | All other Primary Care clinics | 9 | 0%* | 0% | 29.9% |

Table 1 shows the prevalence rates for anti-SARS-CoV-2 total antibody (Ortho Clinical assay) across demographic and clinical covariates of interest. The overall positivity rate for anti-SARS-CoV-2 total antibody was 8.5% (88/1038). SARS-CoV-2 antibodies were found in 8.2% of White children, 5.2% of Black or African American children, 5.7% of Asian children, and 16.2% of children identified as other racial origin. When compared by age groups a bimodal distribution was noted with a seropositive rate of 13.7% in young children 0–5 years, 7.5% in school-age children 6–10 years, 5.1% in early adolescents 11–15 years and 10.8% in older adolescents 16–19 years (Supplemental Figure 1).

Especially high prevalence rates were noted in children with Hispanic ethnicity (26.8%, 55/207), those with public insurance (Medicaid, 21.6% 27/125), those without insurance (55.0%, 11/20), children living in multi-family or apartment dwellings without a private entrance (28.7%, 33/115), and children recruited from safety net primary care clinic (primary care location 2, 25%, 16/64) (Figure 1).

**Figure 1:**
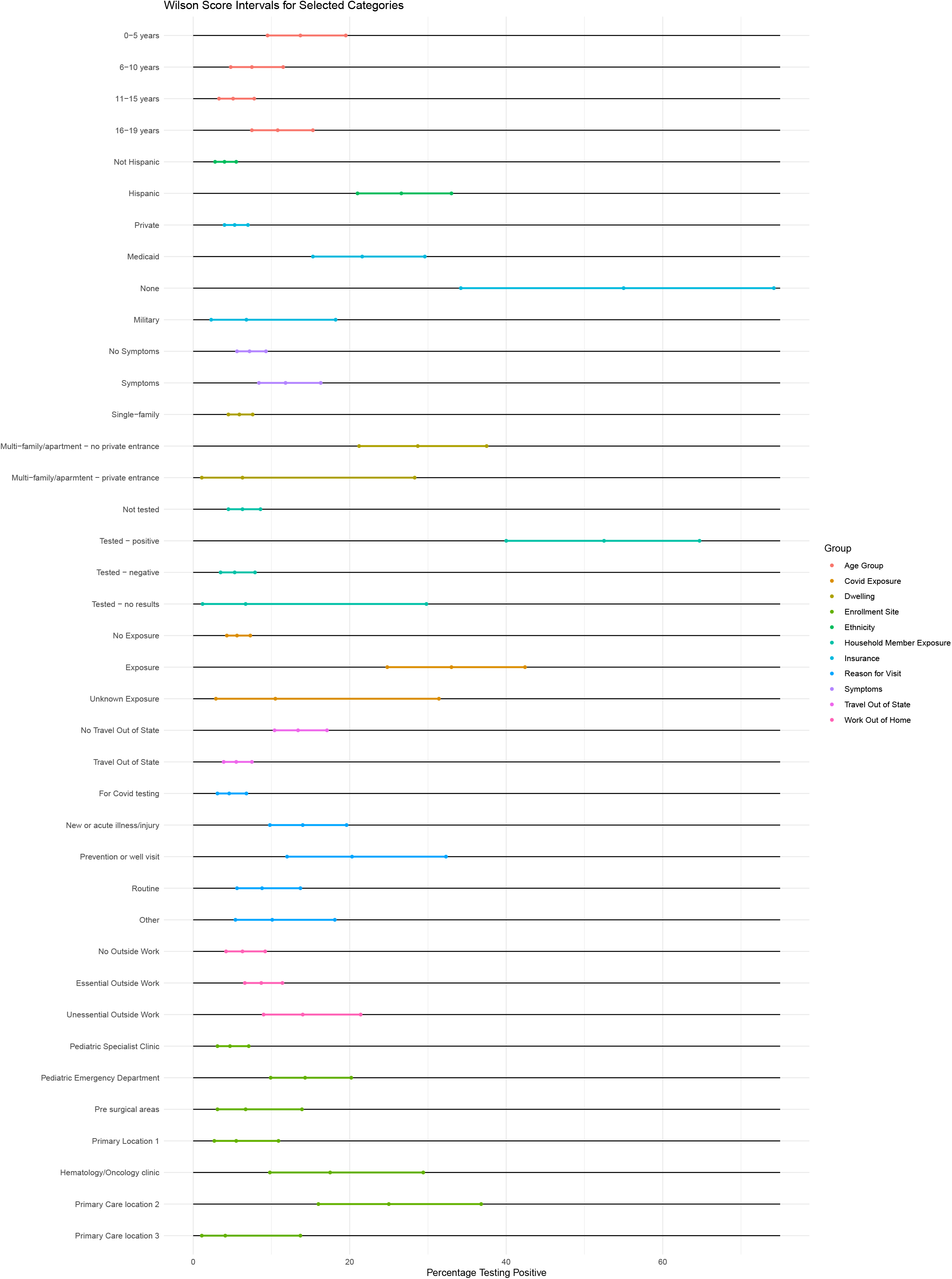
Wilson Score intervals for selected covariates of interest.

Of children exposed to an individual with a known history of COVID-19, 33.0% (35/106) had antibodies. Living in the same household with a person who tested positive for SARS-CoV-2 increased the seroprevalence rate to 52.5% (31/59). Having a personal history of symptoms consistent with COVID-19 was associated with a seroprevalence of 11.8% (30/255). However, 65.9% (58/88) of children with positive antibody testing had no personal history of symptoms, and 54.6% (48/88) had no known exposure.

After multiple regression, significant factors associated with seropositivity were found to include ethnicity, age group, insurance status, symptoms consistent with COVID-19, having a household member test positive for COVID-19 and dwelling type (Table 2).

**Table 2:**
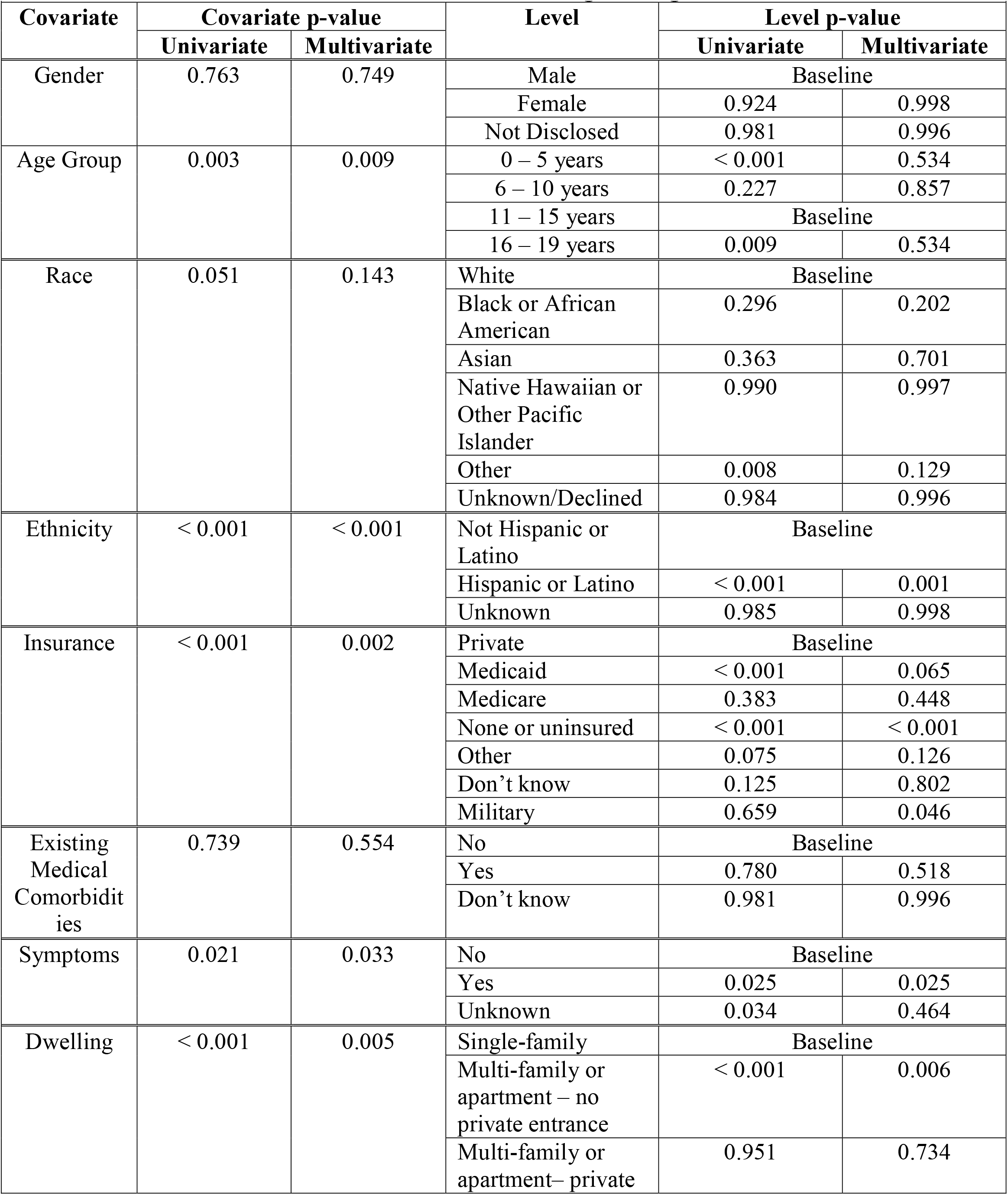
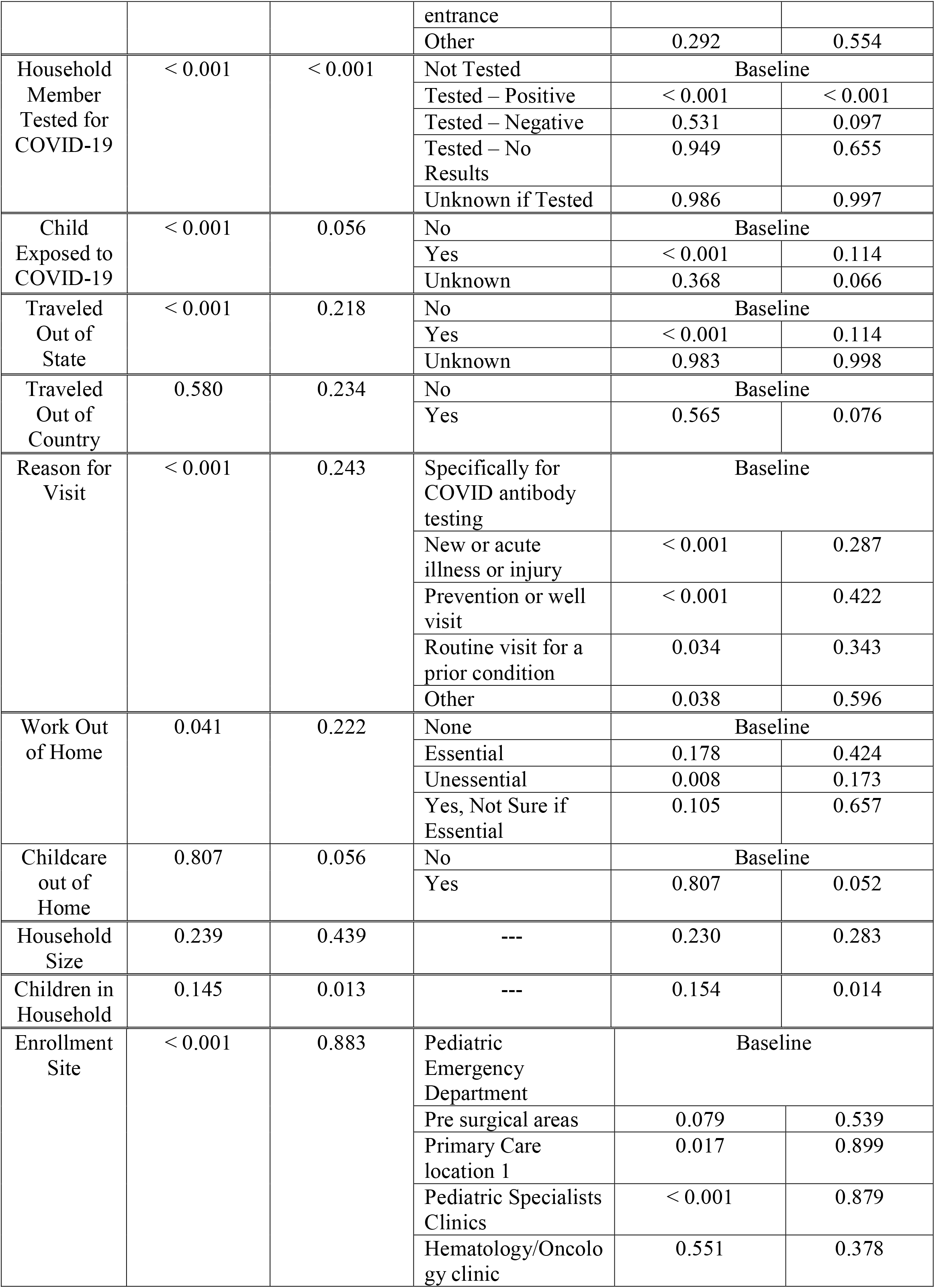
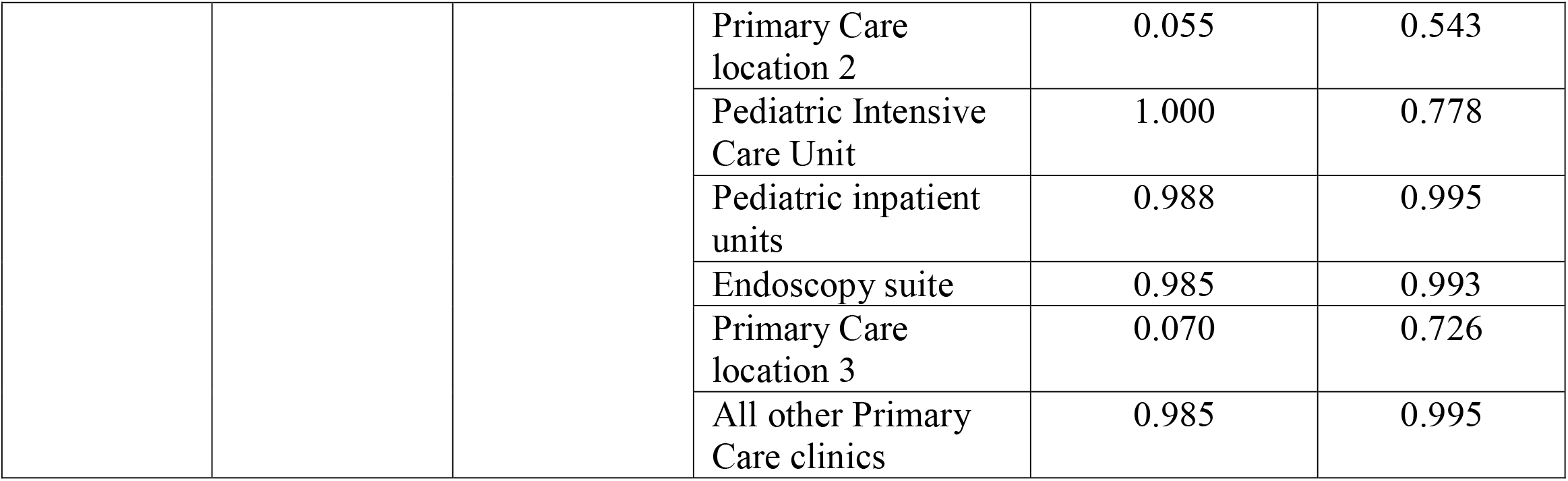
Univariate and Multivariate Logistic Regression Results.

| **Table 2: Univariate and Multivariate Logistic Regression Results** |  |  |  |  |  |
| --- | --- | --- | --- | --- | --- |
| **Covariate** | **Covariate p-value** |  | **Level** | **Level p-value** |  |
|  | **Univariate** | **Multivariate** |  | **Univariate** | **Multivariate** |
| Gender | 0.763 | 0.749 | Male | Baseline |  |
|  |  |  | Female | 0.924 | 0.998 |
|  |  |  | Not Disclosed | 0.981 | 0.996 |
| Age Group | 0.003 | 0.009 | 0 – 5 years | < 0.001 | 0.534 |
|  |  |  | 6 – 10 years | 0.227 | 0.857 |
|  |  |  | 11 – 15 years | Baseline |  |
|  |  |  | 16 – 19 years | 0.009 | 0.534 |
| Race | 0.051 | 0.143 | White | Baseline |  |
|  |  |  | Black or African American | 0.296 | 0.202 |
|  |  |  | Asian | 0.363 | 0.701 |
|  |  |  | Native Hawaiian or Other Pacific Islander | 0.990 | 0.997 |
|  |  |  | Other | 0.008 | 0.129 |
|  |  |  | Unknown/Declined | 0.984 | 0.996 |
| Ethnicity | < 0.001 | < 0.001 | Not Hispanic or Latino | Baseline |  |
|  |  |  | Hispanic or Latino | < 0.001 | 0.001 |
|  |  |  | Unknown | 0.985 | 0.998 |
| Insurance | < 0.001 | 0.002 | Private | Baseline |  |
|  |  |  | Medicaid | < 0.001 | 0.065 |
|  |  |  | Medicare | 0.383 | 0.448 |
|  |  |  | None or uninsured | < 0.001 | < 0.001 |
|  |  |  | Other | 0.075 | 0.126 |
|  |  |  | Don't know | 0.125 | 0.802 |
|  |  |  | Military | 0.659 | 0.046 |
| Existing Medical Comorbidities | 0.739 | 0.554 | No | Baseline |  |
|  |  |  | Yes | 0.780 | 0.518 |
|  |  |  | Don't know | 0.981 | 0.996 |
| Symptoms | 0.021 | 0.033 | No | Baseline |  |
|  |  |  | Yes | 0.025 | 0.025 |
|  |  |  | Unknown | 0.034 | 0.464 |
| Dwelling | < 0.001 | 0.005 | Single-family | Baseline |  |
|  |  |  | Multi-family or apartment – no private entrance | < 0.001 | 0.006 |
|  |  |  | Multi-family or apartment – private entrance | 0.951 | 0.734 |
|  |  |  | Other | 0.292 | 0.554 |
| Household Member Tested for COVID-19 | < 0.001 | < 0.001 | Not Tested | Baseline |  |
|  |  |  | Tested – Positive | < 0.001 | < 0.001 |
|  |  |  | Tested – Negative | 0.531 | 0.097 |
|  |  |  | Tested – No Results | 0.949 | 0.655 |
|  |  |  | Unknown if Tested | 0.986 | 0.997 |
| Child Exposed to COVID-19 | < 0.001 | 0.056 | No | Baseline |  |
|  |  |  | Yes | < 0.001 | 0.114 |
|  |  |  | Unknown | 0.368 | 0.066 |
| Traveled Out of State | < 0.001 | 0.218 | No | Baseline |  |
|  |  |  | Yes | < 0.001 | 0.114 |
|  |  |  | Unknown | 0.983 | 0.998 |
| Traveled Out of Country | 0.580 | 0.234 | No | Baseline |  |
|  |  |  | Yes | 0.565 | 0.076 |
| Reason for Visit | < 0.001 | 0.243 | Specifically for COVID antibody testing | Baseline |  |
|  |  |  | New or acute illness or injury | < 0.001 | 0.287 |
|  |  |  | Prevention or well visit | < 0.001 | 0.422 |
|  |  |  | Routine visit for a prior condition | 0.034 | 0.343 |
|  |  |  | Other | 0.038 | 0.596 |
| Work Out of Home | 0.041 | 0.222 | None | Baseline |  |
|  |  |  | Essential | 0.178 | 0.424 |
|  |  |  | Unessential | 0.008 | 0.173 |
|  |  |  | Yes, Not Sure if Essential | 0.105 | 0.657 |
| Childcare out of Home | 0.807 | 0.056 | No | Baseline |  |
|  |  |  | Yes | 0.807 | 0.052 |
| Household Size | 0.239 | 0.439 | --- | 0.230 | 0.283 |
| Children in Household | 0.145 | 0.013 | --- | 0.154 | 0.014 |
| Enrollment Site | < 0.001 | 0.883 | Pediatric Emergency Department | Baseline |  |
|  |  |  | Pre surgical areas | 0.079 | 0.539 |
|  |  |  | Primary Care location 1 | 0.017 | 0.899 |
|  |  |  | Pediatric Specialists Clinics | < 0.001 | 0.879 |
|  |  |  | Hematology/Oncology clinic | 0.551 | 0.378 |
|  |  |  | Primary Care location 2 | 0.055 | 0.543 |
|  |  |  | Pediatric Intensive Care Unit | 1.000 | 0.778 |
|  |  |  | Pediatric inpatient units | 0.988 | 0.995 |
|  |  |  | Endoscopy suite | 0.985 | 0.993 |
|  |  |  | Primary Care location 3 | 0.070 | 0.726 |
|  |  |  | All other Primary Care clinics | 0.985 | 0.995 |

A final multivariate logistic regression model was chosen using stepwise model selection. Remaining significant predictors included Hispanic ethnicity, lack of insurance or public insurance, a history of symptoms suggestive of COVID-19, exposure to an individual with a history of COVID-19, a household member testing positive for COVID-19, and living in a multi-family or apartment dwelling without private entrance (Table 3).

**Table 3:** Odds Ratio Estimates and p-values for Chosen Covariates.

| **Table 3: Odds Ratio Estimates and p-values for Chosen Covariates** |  |  |  |  |  |
| --- | --- | --- | --- | --- | --- |
| **Covariate** | **Level** | **Univariate Analysis** |  | **Multivariate Analysis** |  |
|  |  | **OR (95% CI)** | **p-value** | **OR (95% CI)** | **p-value** |
| Ethnicity | Not Hispanic or Latino | Baseline |  | Baseline |  |
|  | Hispanic or Latino | 8.73 (5.51, 14.02) | < 0.001 | 4.22 (2.26, 7.94) | < 0.001 |
|  | Unknown | --- | 0.985 | --- | 0.994 |
| Insurance | Private | Baseline |  | Baseline |  |
|  | Medicaid | 4.95 (2.91, 8.31) | < 0.001 | 2.45 (1.10, 5.39) | 0.027 |
|  | Medicare | 2.56 (0.14, 14.86) | 0.383 | 0.47 (0.01, 7.05) | 0.628 |
|  | None or uninsured | 21.94 (8.65, 57.16) | < 0.001 | 11.95 ( 3.71, 38.43) | < 0.001 |
|  | Other | 8.98 (0.41, 95.46) | 0.075 | 5.11 (0.22, 60.29) | 0.206 |
|  | Don't know | 5.98 (0.29, 47.83) | 0.125 | 1.93 ( 0.08, 19.29) | 0.603 |
|  | Military | 1.31 (0.31, 3.80) | 0.659 | 3.37 ( 0.73, 11.40) | 0.075 |
| Symptoms | No | Baseline |  | Baseline |  |
|  | Yes | 1.71 (1.06, 2.71) | 0.025 | 1.99 (1.05, 3.72) | 0.033 |
|  | Unknown | 6.41 (0.88, 33.59) | 0.034 | 4.36 (0.27, 53.50) | 0.292 |
| Dwelling | Single-family | Baseline |  | Baseline |  |
|  | Multi-family – no private entrance | 6.44 (3.92, 10.48) | < 0.001 | 3.19 (1.57, 6.43) | 0.001 |
|  | Multi-family – private entrance | 1.07 (0.06, 5.42) | 0.951 | 0.71 (0.04, 4.63) | 0.765 |
|  | Other | 3.20 (0.17, 20.31) | 0.292 | 1.15 (0.03, 17.95) | 0.931 |
| Household Member COVID-19 Test Results | Not Tested | Baseline |  | Baseline |  |
|  | Tested – Positive | 16.57 (8.98, 31.02) | < 0.001 | 9.65 (3.34, 30.55) | < 0.001 |
|  | Tested – Negative | 0.84 (0.48, 1.45) | 0.531 | 0.69 (0.35, 1.33) | 0.281 |
|  | Tested – No Results | 1.07 (0.06, 5.57) | 0.949 | 2.36 (0.12, 15.00) | 0.446 |
|  | Unknown if Tested | --- | 0.986 | --- | 0.993 |
| Child Exposed to COVID-19 | No | Baseline |  | Baseline |  |
|  | Yes | 8.31 (5.06, 13.60) | < 0.001 | 2.93 (1.06, 7.45) | 0.030 |
|  | Unknown | 1.98 (0.31,<br>7.18) | 0.368 | 3.22 (0.46,<br>13.79) | 0.160 |
| Travel Out of<br>State | No | Baseline |  | Baseline |  |
|  | Yes | 0.37 (0.24,<br>0.58) | < 0.001 | 0.53 (0.28,<br>0.98) | 0.045 |
|  | Unknown | --- | 0.983 | --- | 0.995 |
| Number of Children |  | 0.84 (0.66,<br>1.06) | 0.154 | 0.67 (0.48,<br>0.92) | 0.016 |

Of those who tested positive for anti-SARS-CoV-2, titer levels ranged from 1.05 to 1000 S/Co. Across all age groups, titer levels did not significantly differ (Kruskal-Wallis, p=0.088) (Supplemental Figure 2). There was no significant difference in titer levels between those with no symptoms, those with symptoms within 2 weeks of testing, and those with symptoms more than 2 weeks before testing (p-value=0.893, Kruskal-Wallis) (Supplemental Figure 3).

Orthogonal testing was performed on 87/88 reactive samples (where enough serum remained) using the Siemens assay against the S1/RBD target. This demonstrated an 80.5% agreement with 70/87 samples reactive. Those with lower titer levels on the Ortho Clinical assay were more likely to test negative on the Siemens assay (p=0.001, logistic regression) and older individuals were more likely to test negative (p=0.011, logistic regression); Figure 2a. There was no significant different between those with recent symptoms within 2 weeks versus those with more remote symptoms in negative results on orthogonal testing.

**Figure 2.**
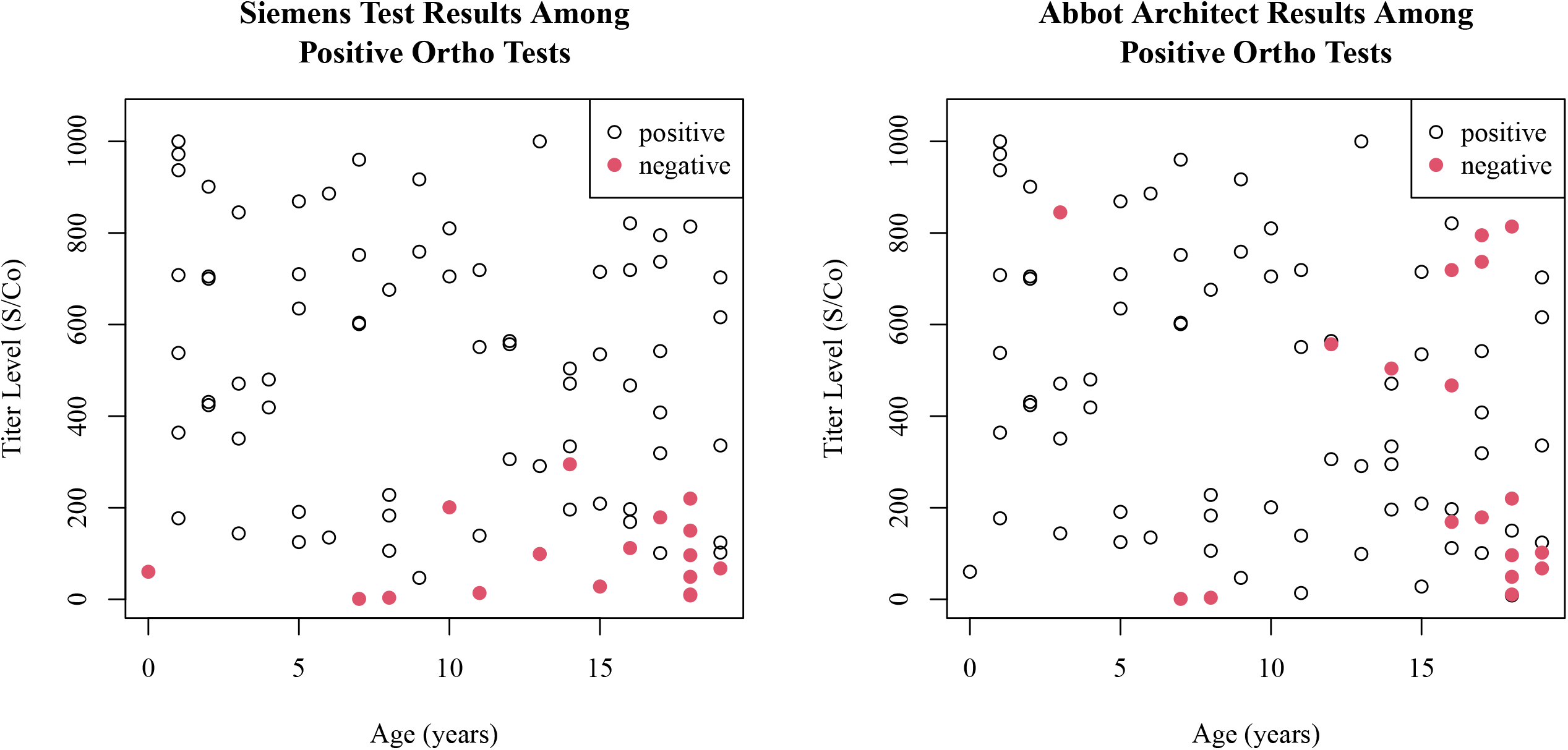
Scatterplots showing age versus titer levels of the reactive tests on the Ortho Clinical Diagnostic Vitros assay, with orthogonal testing on the Siemens assay (2a) and Abbot assay (2b). Negative tests on orthogonal testing are colored red.

When comparing reactive samples from the Ortho Clinical assay with the Abbot assay targeting the nucleopcapsid antigen there was a 79.3% agreement with 69/87 samples testing positive. Those in older age groups were more likely to test negative on the Abbot assay (p=0.001, logistic regression) but there was no significant change among titer levels (p=0.177, logistic regression); Figure 2b.

A random selection of 98 samples (approximately 10%) that tested non-reactive on the Ortho Clinical assay were tested on both the Abbot and Siemens assays. All samples were negative on the additional two assays.

## Discussion

Using the Ortho Clinical anti-SARS-CoV-2 antibody assay, it was determined that the seroprevalence of SARS-CoV-2 in children in Northern Virginia was as high as 8.5% (95% CI 6.9-10.3) in the fall of 2020. In contrast, the concurrently reported prevalence of SARS-CoV-2 in children in Virginia and throughout the US, based on molecular and antigen tests at the time of study completion, was around 1% [11]. Our seropositive rate of infection in children was almost 8 times the reported prevalence based on real-time viral testing.

An independently determined Northern Virginia adult seroprevalence was 4.4% in a similar study that concluded 2 months prior to this pediatric study (personal communication, author Christopher Difillipi). As of mid-October, the Centers for Disease Control reported a Virginia adult seroprevalence of 4.1% [12]. The seropositivity rate in children in this current study was more than double the reported concurrent adult seroprevalence.

The higher than expected pediatric seropositivity rate in our study is in part likely attributable to continued viral transmission over time. In addition, early in the pandemic, it was perceived that children were at reduced risk for acquiring COVID-19, either from innate protective physiologic characteristics or reduced exposure [1,3]. Schools closed rapidly at the start of the pandemic with reduced communal exposure. However, the unrecognized burden of mild or asymptomatic disease, resulted in testing bias and under-representation of children in early testing schemes [1, 13-15]. The high SARS-CoV-2 seropositivity in children we identified was an unexpected finding and it has important implications for this silent burden of disease and risk of transmission to and from children.

In our population, after multi-regression analysis, identified significant risk factors for seropositivity included: age-group, Hispanic ethnicity, insurance status, and residence within a multi-family dwelling or apartment without a private entrance. The highest risk belonged to children living in the same home as an individual who had tested positive for SARS-CoV-2, with an odds ratio of 22.

Children under 5 years were statistically more likely to be seropositive for SARS-CoV-2 compared with other age groups in univariate analysis. This finding was also surprising and has implications for the need for different considerations in mitigation and protection strategies in the younger age groups. Neither the Spanish nor Switzerland cohorts examined children <5 years, but adolescents (ages 10-19) in Switzerland were more likely to test positive for antibodies than younger children (ages 5-9) [3,4]. Young children are cared for more closely by adults with possible higher risk of exposure due to prolonged physical proximity and are more likely to interact with each other with less protection. Additionally, even though there was an indoor mask mandate outside the home, this was only for children ≥10 years. Moreover, even as public schools were closed for in-person learning, some childcare facilities for younger children remained open. However, consistent with other reports, our analyses did not indicate a significant difference in seropositivity in children cared for outside of their home, probably because greater efforts are taken to prevent transmission in childcare settings [16].

The majority of children with anti-SARS-CoV-2 antibodies in this cohort (66%) did not recall having symptoms of COVID-19. While the veracity of this is affected by recall bias, this highlights the high proportion of infections that are asymptomatic in the pediatric population and the subsequent risk of transmission to other vulnerable persons.

For seropositive individuals, mean antibody titers did not appear to differ between those with or without symptoms, recent or remote. It is interesting that for some seropositive children, symptoms were reported <2 weeks from testing as the reported kinetics of the antibody response to SARS-CoV-2 demonstrate that at least a week is necessary to mount IgM responses after onset of symptomatic disease [15]. Since COVID-19 symptoms overlap greatly with other childhood viral syndromes, it is possible that these reported symptoms were not truly caused by COVID-19 infection. It is also possible that some children with remote symptomatology had seroreversion and so were not captured in the positive cohort [17,18].

Exposure to infectious persons with COVID-19 is a well-recognized risk factor for acquiring COVID-19 and our analysis confirms this [19-22]. Consistent with other reports, the household attack rate was even higher; over 50% of children reported as exposed to a test-positive household member were seropositive [23]. Conversely, almost 60% of seropositive children had no known contact with test-positive SARS-CoV-2. This emphasizes the insidious effects of asymptomatic spread with viral shedding in symptomatic and asymptomatic individuals [24].

Ethnically and socioeconomically marginalized groups unable to shelter in place have shouldered a disproportionate burden of COVID-19 disease globally [23,26,27]. Hispanic ethnicity, the dominant minority in Northern Virginia, conferred four-fold odds of seropositivity. Publicly provided health insurance and lack of insurance is an accepted marker for lower socioeconomic status, and families with limited financial means are more likely to live in multi-family dwellings with a common entrance. Significantly higher seropositivity rates in each of these groups demonstrated the disproportionate impact of COVID-19 on the underprivileged.

Orthogonal testing with an antibody assay specific to the S1/RBD antigen of the spike protein demonstrated high concordance. Those samples which failed to show concordance in orthogonal testing to the S1/RBD exhibited very low titers to the full spike protein (Ortho Clinical assay). Future consideration should be given to redefining the lower limit of detection for full spike protein antibody assays as it is possible that these lower titer levels may represent cross-reactivity to conserved epitopes in benign seasonal human coronaviruses [8]. In addition, some samples that may have been negative on orthogonal testing may have been due to the Ortho Clinical assay also detecting IgM; however those negative on orthogonal testing were not more likely to have recent symptoms consistent with COVID-19 within 2 weeks of antibody testing.

Children with a response to the full spike protein assay also frequently manifested antibodies to the nucleocapsid antigen. Interestingly, it was those in older age groups that more likely failed to produce a nucleocapsid antibody response, contrary to previously published data suggesting that young children may fail to elicit a nucleocapsid antibody response [7].

## Limitations

There are several potential limitations to this investigation. Selection bias may have affected the representativeness of the regional population, as it was focused on children having blood drawn for another clinical purpose and self-referral for the study. However, these factors are accounted for in this analysis. In addition, Northern Virginia is a major metropolitan area, and so results may not be generalizable to areas that have more rural demographics. As noted, cross-reaction with other common endemic human beta-coronaviruses may result in false positive antibody results, particularly at currently defined thresholds [8]. Conversely, seroreversion may produce false negative results in more remote infections [23,24]. Most importantly, this study took place at a specific point in time and represents a static snapshot of a dynamic event [4].

## Conclusions

A much higher pediatric SARS-CoV-2 burden was found than has previously been reported in the US; this rate was also higher than in adults in the same region over a similar time period. These seropositive rates were following the period after the first peak of infection in the community, when schools were closed, and social mitigation strategies were mandated. Contrary to reporting early in the COVID-19 pandemic, this study determined that children experience a significant burden of COVID-19 infection. Children appear very important in the transmission of disease, including silent transmission, and with such continued protection of children through social mitigation strategies and extending COVID-19 vaccines to children are imperative to achieving control of this pandemic.

## Data Availability

Data sharing requests will be considered by the authors upon written request to the corresponding author. Deidentified participant data or other prespecified data will be available subject to a written proposal and a signed data sharing agreement.

## Acknowledgements

We recognize the efforts of all team members at the multiple study sites who have supported this study and encouraged participation. This project would not have been possible without the support of Dr. David Trump and the Virginia Department of Health, the help of Dr. John Farrell and Dr. Dina Gottsman and the pediatric team at Farrell Pediatrics, Dr. Albert Brito and the pediatric team at Inova Cares Clinic for Children, Dr. Sandy Chung and the pediatric team at Fairfax Pediatrics, and Dr. Christina Ulen, Ms. Lori Schubert, and the pediatric team at Advanced Pediatrics, Dr. Elizabeth Yang and the pediatric teams at Pediatric Specialists of Virginia, the pediatric teams at Inova Children’s Hospital. We recognize and thank all of the participants of the study and their families; such studies can only happen because people find that helping in research, especially in the midst of a pandemic, is worthwhile, and therefore, choose to join.

## Figure Legends

Supplemental Figure 1: Pairwise plot of age vs. seropositivity rate (%). The red points represent the observed positivity rate in the cohort at each age, and the line is the predicted positivity rate produced by a quadratic regression model. The estimated seropositivity rate is higher for younger and older ages of children.

Supplemental figure 2: A multiple boxplot that shows the SARS-CoV-2 total spike protein titer levels (S/Co) for those who tested positive separated by age group and summary statistics.

Supplemental Figure 3: A multiple boxplot that showing SARS-CoV-2 total spike protein titer levels (S/Co) for those who tested positive separated by presence of symptoms and summary statistics.

