## Supplementary figures and images for "SARS-CoV-2 Seroepidemiology in Children and Adolescents"

### Supplementary Figure 1

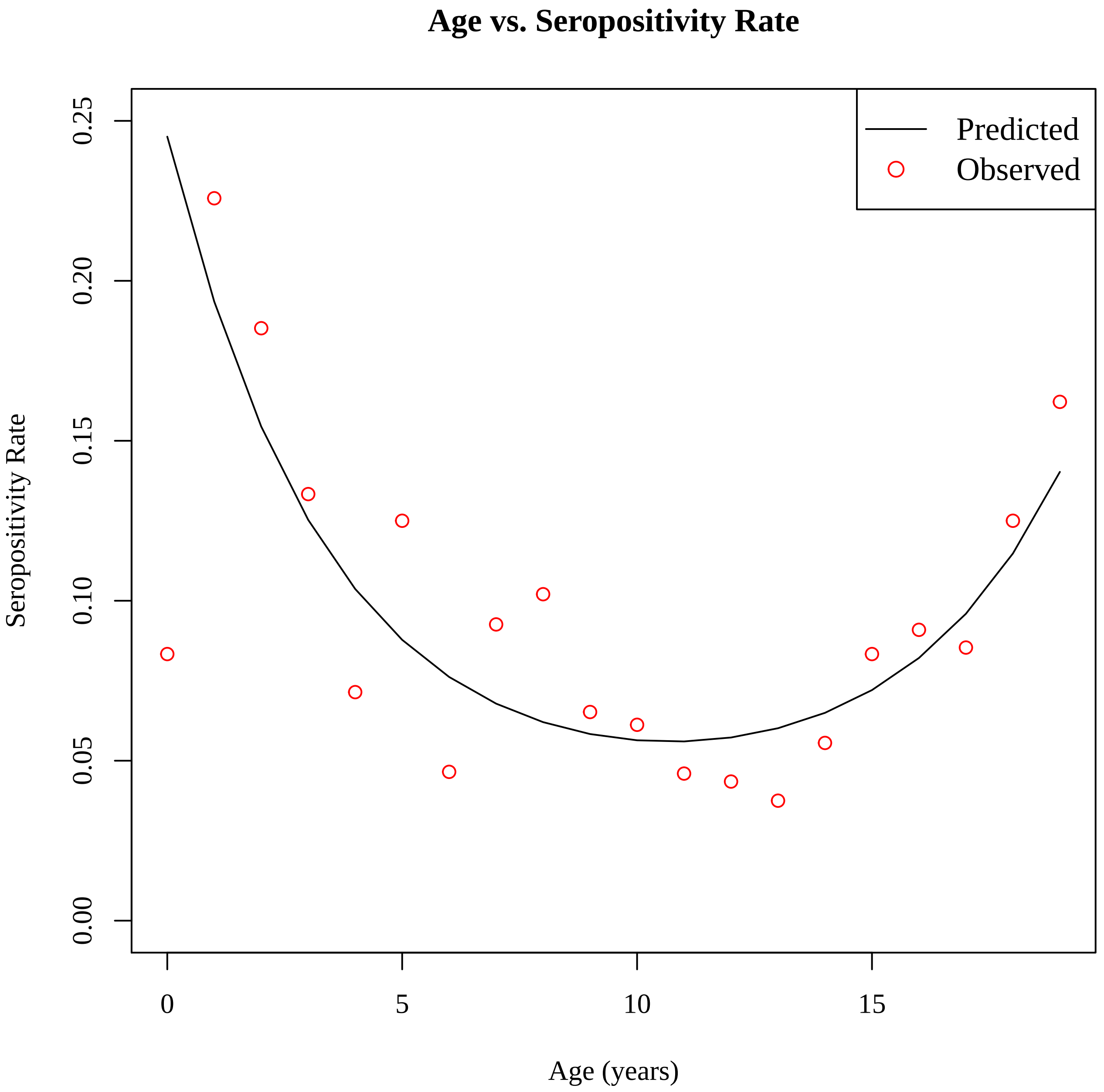

### Supplementary Figure 3

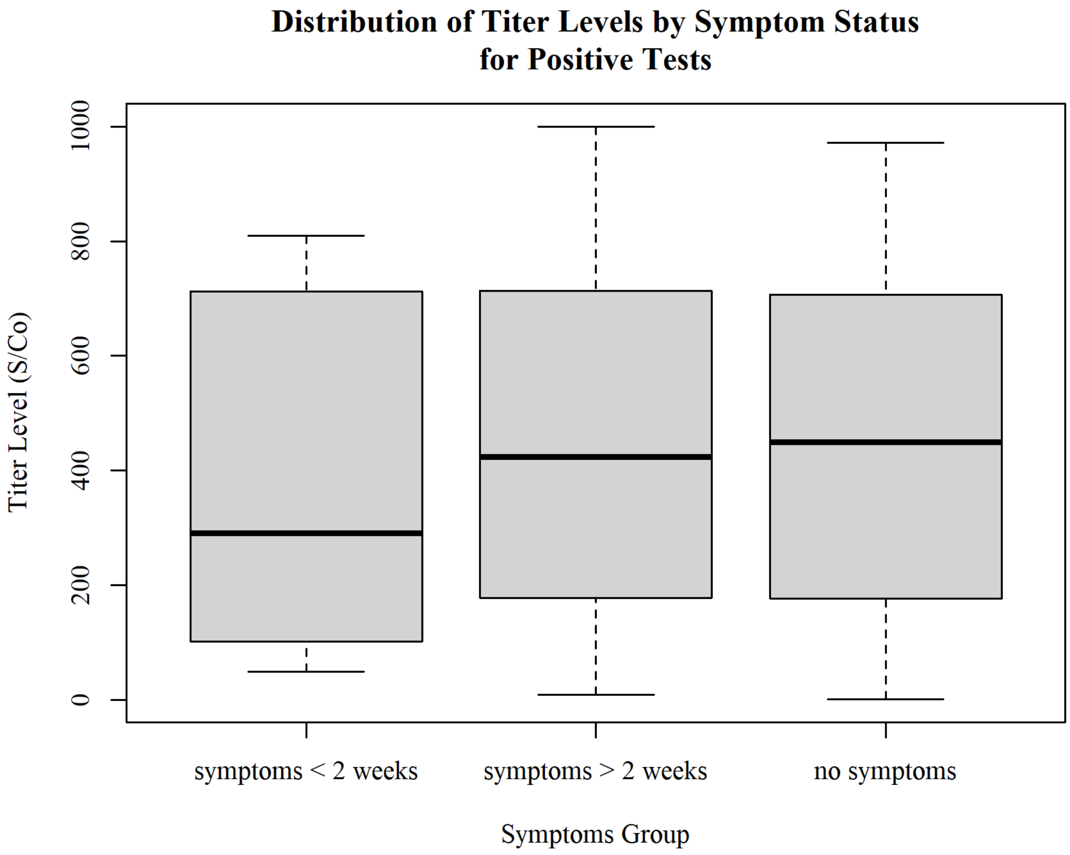


| Group | Count | Mean | Std. Dev. | Median | IQR |
|---|---|---|---|---|---|
| symptoms < 2 weeks | 7 | 297 | 336 | 291 | 610 |
| symptoms > 2 weeks | 23 | 460 | 323 | 424 | 535 |
| no symptoms | 56 | 443 | 295 | 449 | 526 |
