## Supplementary Figure 2 for "SARS-CoV-2 Seroepidemiology in Children and Adolescents"

### Distribution of Titer Levels by Age Group for Positive Tests

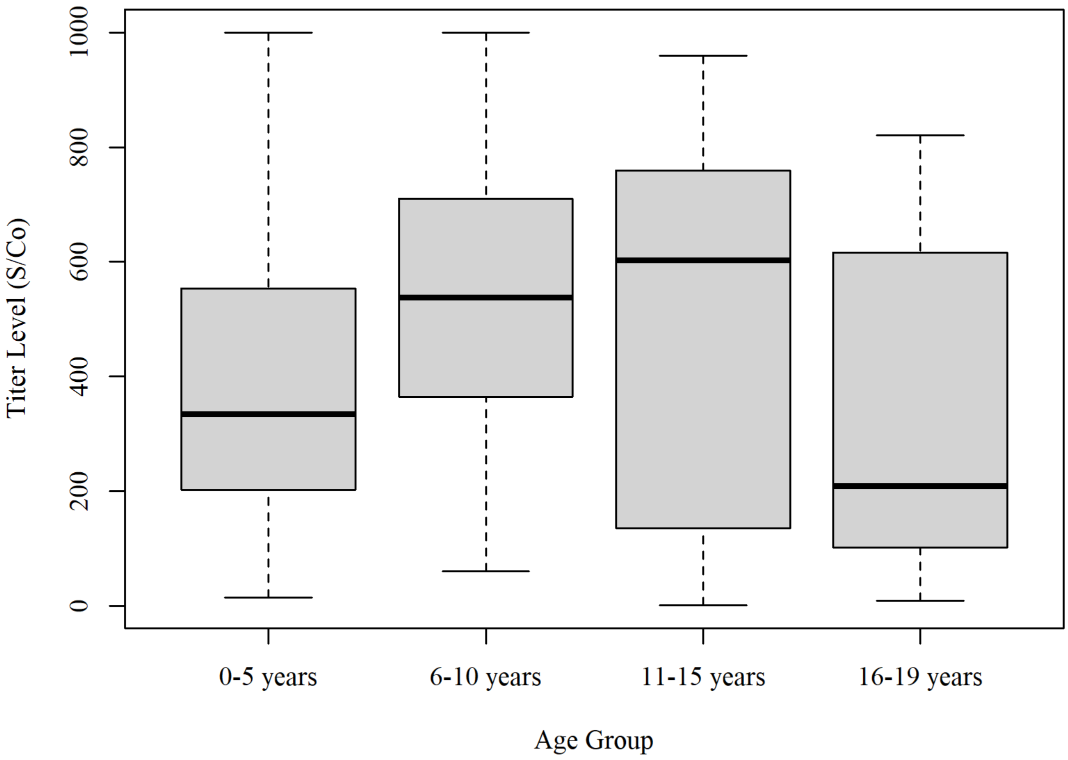


| Group | Count | Mean | Std. Dev. | Median | IQR |
| --- | --- | --- | --- | --- | --- |
| 0-5 years | 25 | 553 | 285 | 538 | 346 |
| 6-10 years | 18 | 476 | 351 | 602 | 610 |
| 11-15 years | 19 | 396 | 261 | 334 | 352 |
| 16-19 years | 26 | 341 | 284 | 208 | 493 |
